# Monitoring sick leave data for early detection of influenza outbreaks

**DOI:** 10.1101/2020.05.28.20115782

**Authors:** Tom Duchemin, Jonathan Bastard, Pearl Anne Ante-Testard, Rania Assab, Oumou Salama Daouda, Audrey Duval, Jérôme-Philippe Garsi, Radowan Lounissi, Narimane Nekkab, Helene Neynaud, David R. M. Smith, William Dab, Kevin Jean, Laura Temime, Mounia N. Hocine

**Affiliations:** MESuRS laboratory, Conservatoire National des Arts et Métiers, 292 Rue Saint-Martin, 75003 Paris, France; Malakoff Humanis, 21 Rue Laffitte, 75009 Paris, France; Institut Pasteur, Epidemiology and Modelling of Antibiotic Evasion (EMAE), Paris, France; PACRI unit, Conservatoire National des Arts et Métiers, Institut Pasteur, Paris, France; Université Paris-Saclay, UVSQ, Inserm, CESP, Anti-infective evasion and pharmacoepidemiology team, Montigny-Le-Bretonneux, France; Malaria: Parasites and Hosts, Department of Parasites and Insect Vectors, Institut Pasteur, Paris, France; Biodiversity and Epidemiology of Bacterial Pathogens, Institut Pasteur, Paris, France

## Abstract

**Background:** Workplace absenteeism increases significantly during influenza epidemics. Sick leave records may facilitate more timely detection of influenza outbreaks, as trends in increased sick leave may precede alerts issued by sentinel surveillance systems by days or weeks. Sick leave data have not been comprehensively evaluated in comparison to traditional surveillance methods.

**Aim:** To study the performance and the feasibility of using a detection system based on sick leave data to detect influenza outbreaks

**Methods:** Sick leave records were extracted from private French health insurance data, covering on average 209,932 companies per year across a wide range of sizes and sectors. We used linear regression to estimate the weekly number of new sick leave spells from 2010 to 2017 in 12 French regions, adjusting for trend, seasonality and worker leaves. Outbreaks were detected using a 95%-prediction interval. This method was compared to results from the French Sentinelles network, a gold-standard primary care surveillance system currently in place.

**Results:** Using sick leave data, we detected 92% of reported influenza outbreaks between 2016 and 2017, on average 5.88 weeks prior to outbreak peaks. Compared to the existing Sentinelles model, our method had high sensitivity (89%) and specificity (86%), and detected outbreaks on average 2.5 weeks earlier.

**Conclusion:** Sick leave surveillance could be a sensitive, specific and timely tool for detection of influenza outbreaks.

## 1. Introduction

Early outbreak detection is crucial for preparedness and timely public health and medical responses. It provides useful information to physicians, companies, and the public, ensuring proper drug prescription, health service planning, workplace preparedness, and continuity of operations in case of high absenteeism (1), among many other uses.

Most countries face periodic influenza (or “flu”) epidemics that vary in size and severity from year to year (2). Seasonal flu can be highly virulent and, like many respiratory viruses, can spread rapidly through populations highlighting a need for a robust epidemiological surveillance system to detect emerging outbreaks. Surveillance system guidelines developed by the US Centers for Disease Control (CDC) suggest that systems should be simple, reliable, flexible, timely, and readily accepted by diverse individuals and organizations to ensure participation (3).

Flu surveillance systems vary by country and rely on various types of data. National health agencies monitor flu epidemics using healthcare records, medical sentinel systems, pharmaceutical sales and other data sources. Most of these systems rely on data from healthcare settings that rely on patient healthcare seeking behavior or after results of clinical tests, which often reflects those with symptoms or relatively advanced stages of the disease. These systems fail to capture individuals who do not seek medical care, whether due to asymptomatic infection, perceived mildness of infection or a general reluctance to seek care (1,4). Given these gaps, alternative data streams from non-healthcare settings may provide a valuable complement to classical surveillance systems (5).

Human resources data collected at the workplace have received relatively little attention for outbreak detection, but present characteristics that are useful for infectious disease surveillance. In many settings, absenteeism data are routinely collected and centralized either for the use by companies themselves or for health insurance purposes. Due to legal purposes and to the implication of work absence on salaries, these data are comprehensive and reliable. Though a handful of studies have assessed the role of sick leave data for outbreak detection, they did not develop a comprehensive assessment of its performance and robustness. Bollaerts et al., Patterson et al. and Groenewold et al. (1,6,7) assessed the usefulness of work absenteeism surveillance as a tool for early warning systems for influenza. They can also supplement more traditional medical data by providing information about an epidemic’s socioeconomic impact (1,4). As a consequence, the US National Institute for Occupational Safety and Health (NIOSH) has been monitoring health-related workplace absenteeism among full-time workers using data received monthly from the Current Population Survey since 2017 and making this data available online (8).

A great challenge in epidemiological surveillance lies in identifying data streams that allow for sensitive, specific and timely outbreak detection. In the context of outbreak detection, surveillance sensitivity can refer to both (i) the proportion of true cases detected, and (ii) the probability of detecting an outbreak, including the changes in the number of cases over time (3). Surveillance specificity refers to the probability of correctly identifying when an outbreak is not occurring (9). Lastly, surveillance timeliness generally refers to the time difference between an event and its standard reference(5). Some studies have suggested that absenteeism data along with others such as over-the-counter pharmaceutical sales and emergency visits seem to be more timely than sentinel Influenza-Like Illness (ILI) surveillance (5,6), other traditional flu data sources (7), physician diagnoses (5), and virological data (5).

In this study, we assess the sensitivity, specificity and timeliness of workplace absenteeism data for detection of flu outbreaks in France. Our hypothesis is that monitoring sick-leave data at the workplace might help anticipate outbreaks in a timely manner using routinely collected data. We then compare the performance of a sick-leave based monitoring system to the performance of the national standard surveillance system of influenza in France which is based on ILI data.

## 2. Material and methods

### 2.1. Data

#### 2.1.1 Sick-leave data

The study relies on the sick leave record system of the French health insurance company Malakoff Médéric. Malakoff Médéric insures sick leaves for 114,707 to 245,973 French companies in a wide range of sectors, covering between 290,056 and 2,765,400 employees per year. This wide variation is due to the fact that some companies were no longer required to report these data to the insurer after 2015. The companies have in average 18.7 employees per year. Nearly half of companies (46%) were in Services, 36% in Commerce, 12% in Industry and Construction, and 5% in Health.

These data are routinely collected and annually reported in the system DADS (*Déclarations Annuelles de Données Sociales*). For our purposes, we used the weekly incidence rate of sick leave spells (per 100,000 workers) aggregated at the regional level across the 12 administrative regions of metropolitan France over the period 2010–2017.

#### 2.1.2 Workers leave data

The number of workers on non-sick leave (e.g. paid holiday) is not reported in DADS, so the denominator of the weekly sick leave incidence rate was defined as the number of workers actively employed by their company during the observed week, and not the number of workers actually working during the week. To adjust our data, we used data from statistics department of the French Ministry of labour (DARES) to build an indicator (the worker-leave-peak indicator) describing weeks with a peak in sick leave (10). Peaks were identified during the Christmas school holidays (last week of December and first week of January) and during summer (second week of July to third week of August).

#### 2.1.3 Influenza-like illness data

Weekly sick leave incidence was compared to weekly ILI incidence (per 100,000 inhabitants), derived from the French influenza surveillance database of the GP Sentinelles network, coordinated by Santé Publique France. In 2018, the Sentinelles network was composed of 1,314 general private practitioners and 116 private pediatricians, all voluntary participants and spread widely across the whole of France’s territories. Detailed information on this network can be found elsewhere (11). This ILI incidence data is the main source of data used to declare influenza epidemics in France. ILI are defined by Sentinelles as a fever above 39°C, with sudden onset, accompanied by myalgia and respiratory signs.

In addition to providing weekly ILI incidence data, the French Sentinelles network also proposes an ILI-outbreak detection algorithm. The algorithm is based on a Serfling method (12). It has been adapted for routine surveillance of epidemics of ILI in France (13). The method implemented by Sentinelles is based on a periodic regression model including a biannual seasonal effect of clinical influenza, a linear trend and an intercept adjusting for a baseline diagnostic activity (which corresponds to the number of influenza syndromes that would be diagnosed in the absence of influenza virus during the off-season).

### 2.2. Methods

#### 2.2.1 Identification of influenza outbreak episodes

Dates of influenza outbreak episodes from Sentinelles are publicly unavailable so we trained the model described above on data from 1984 to 2009 to mimic the sentinel system. Weekly ILI incidence rates per 100,000 residents and per region from 2010 to 2017 were then compared to an outbreak detection threshold. This was defined as the upper bound of the 95% prediction interval from this model, and to increase specificity an alert was only declared when this threshold was crossed twice consecutively.

#### 2.2.2 Determination of sick leave outbreak episodes

To detect sick leave outbreak episodes, an algorithm based on the Serfling method was used. The regression includes an intercept to adjust for the baseline sick-leave activity and the worker-leave-peak indicator to adjust for seasonality.

Similarly to the Sentinelles method, an outbreak was declared if the true sick leave incidence rate crossed the 95% prediction interval twice consecutively. An alert was lifted when the incidence fell below the threshold, again for two consecutive weeks (14).

The model was trained on sick leave data from 2010 to 2015. Years 2016 and 2017 were used to evaluate model performance. As timing of ILI outbreaks may vary geographically, analyses were conducted separately for each of mainland France’s 12 administrative regions. For simplicity, some results were plotted in the main text for three regions only, chosen to reflect a North-South gradient (respectively Haut-de-France, Ile-de-France and Provence-Alpes-Côte d’Azur). Full results are included as supplementary results.

#### 2.2.3 Criteria for assessing the proposed surveillance system

Evaluation criteria were selected to answer two questions: (i) Does the sick leave model efficiently detect ILI outbreaks? and (ii) How does it compare to the Sentinelles model? Results are presented for each French administrative region and are also aggregated at the national level.

##### Performance of the sick leave model to detect ILI outbreaks

To answer the first question, we calculated sensitivity and specificity per outbreak episode to evaluate whether our model correctly detected all ILI outbreaks. The two criteria are:

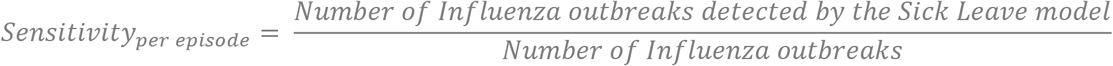

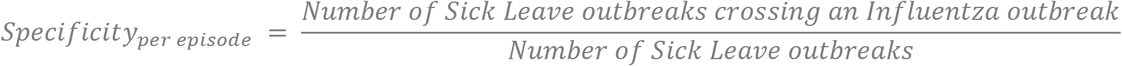

To evaluate our model’s timeliness, we calculated the detection time we defined as the delay between the outbreak detection of our algorithm compared to the annual influenza outbreak peak. The influenza outbreak peak was defined as the week with the highest number of reported ILI cases between June 1st and May 31st of the subsequent year.

##### Performance of the sick leave model compared to the Sentinelles model

To evaluate the performance of the sick leave model, we calculated its sensitivity and specificity with respect to the Sentinelles model. Unlike the previous criteria, we define these indicators at the level of the week rather than the episode. The objective is to assess whether our two models are similar. The two criteria are defined as follows:

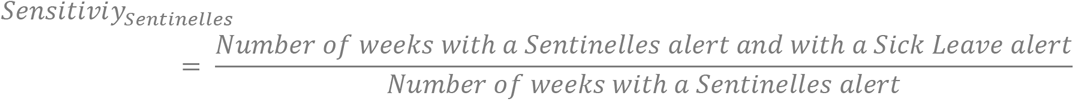

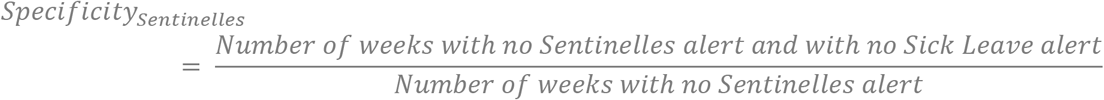

The timeliness of the sick-leave model compared to the Sentinelles model was also evaluated: the delay between the outbreak detection of the first model compared to the second one is computed.

#### 3. Results

Incidence curves obtained from the Sentinelles surveillance networks from 2010 to 2017 reveal annual peaks of influenza-like illness (ILI), from 163 to 1,290 per 100,000 per week, occurring approximately between December and February (Figures 1 and S1). During the summers, the incidence approaches zero. By comparison, weekly sick leave incidence varied about an annual average of 1,021 to 1,335 per 100,000 per week, depending on the region (Figures 1 and S1). They exhibit greater variability and more peaks per year than ILI incidence. However, based on visual inspection, the highest seasonal peaks tend to coincide with ILI incidence peaks. Moreover, most of the seasonal sick-leave troughs coincide with Christmas and summer school holidays periods, which can be explained by a decrease of the at-risk population, i.e. an increase in the number of workers on paid leave. Finally, there is no apparent change between 2014 and 2015 despite the strong variation in the volume of workers in the database.

**Figure 1:**
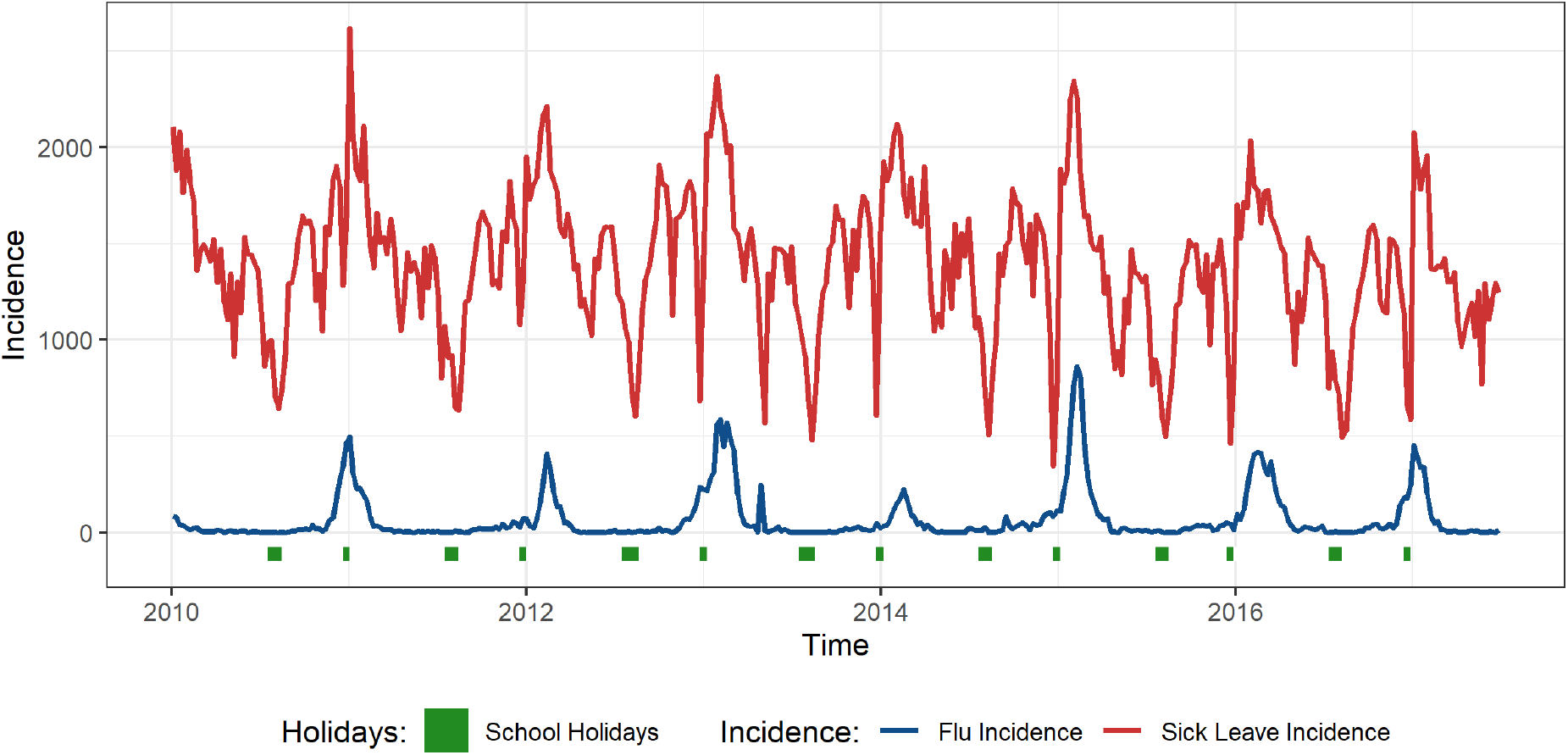
Incidence per 100,000 per week of influenza-like illness and sick leave in Ile-de-France, the most populous region in France, 2010–2017. Christmas and summer school holidays (increased worker leave periods) are shown at the bottom.

For three French regions, Figure 2 presents the incidence of sick leaves and ILI for the 2015–2017 time period. In each region, the Sentinelles surveillance system identified exactly one alert per year, triggered a few weeks before or during the peak of ILI incidence (Figures 2 and S2). The exception was Bretagne, where no alert was identified during winter 2016–2017 (Figure S2). By comparison, the sick leave surveillance system triggered one to three alert episodes per year.

**Figure 2:**
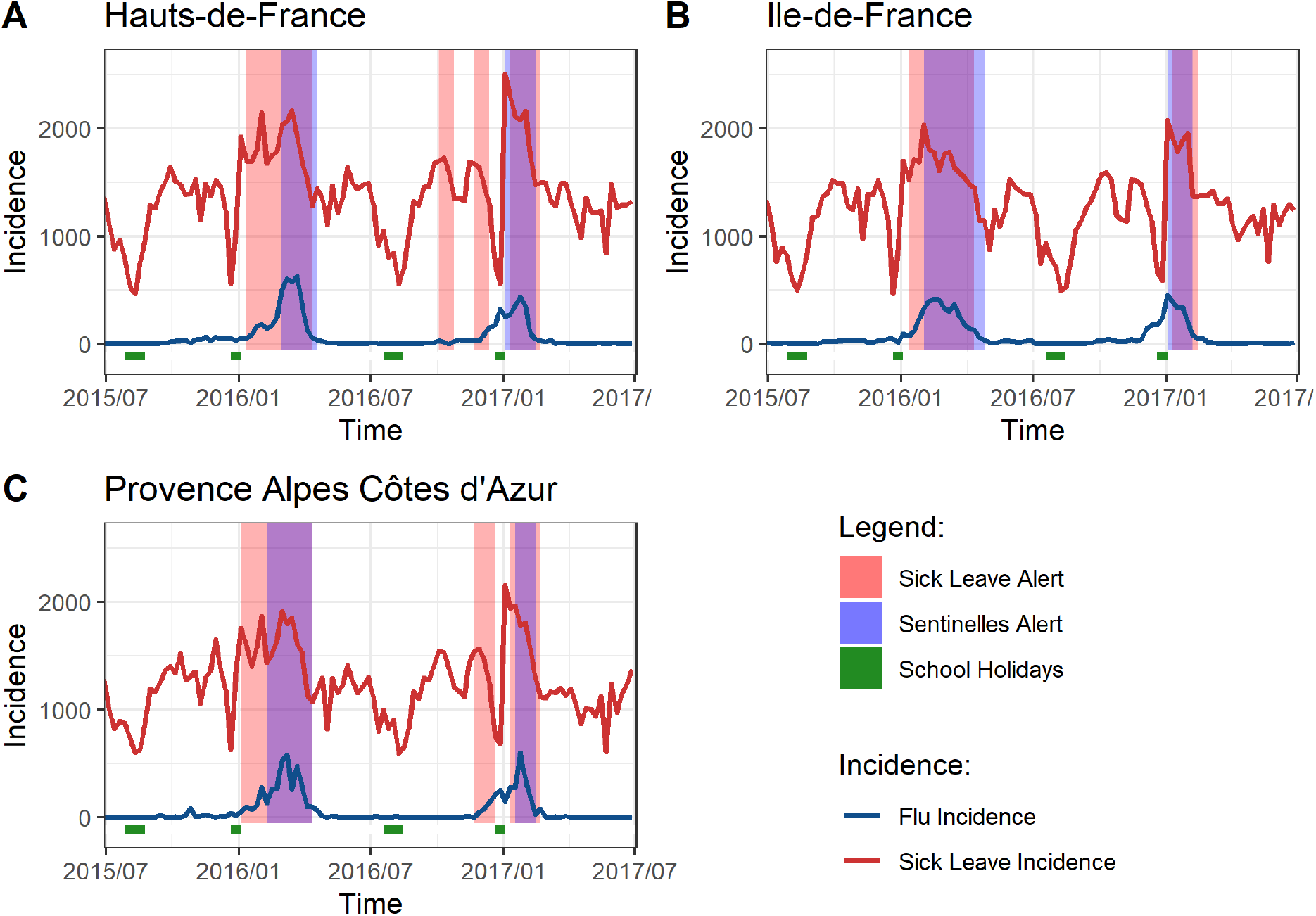
Incidence per 100,000 per week of influenza-like illness and sick leave, 2015–2017, and alerts from the Sentinelles (blue) and the sick-leave (red) models, in three French regions (A: Hautsde-France, B: Ile-de-France, C: Provence-Alpes-Côte d’Azur). Alerts show as purple when Sentinelles and sick-leave alerts overlap. Christmas and summer school holidays (increased worker leave periods) are shown at the bottom (green).

We assessed the sick leave surveillance system on (i) its ability to detect and anticipate ILI incidence peaks, and (ii) how it compared with the Sentinelles surveillance system. Table 1 summarizes the indicators of its performance regarding ILI peak detection and anticipation in each region, averaged over the two years of the model test. The sensitivity per episode (probability of detection of the ILI outbreak) had a mean of 0.92 (range 0.5–1) across regions, while the specificity per episode had an average of 0.58 (range 0.2–1). The sick leave alert generally occured prior to peak ILI incidence, on average 5.88 weeks (range 2.5–11) before the peak.

**Table 1:** Performance of the sick-leave model to detect ILI outbreaks: sensitivity per episode (probability of detection), specificity per episode and detection time before ILI peak. For each region, the value of these indicators are averaged over the two years evaluated.

| Region | Sensitivity per episode | Specificity per episode | Detection time (weeks) before ILI |
| --- | --- | --- | --- |

|  |  |  | **peak** |
| --- | --- | --- | --- |
| **Auvergne-Rhône-Alpes** | 0.5 | 0.2 | 7 |
| **Bourgogne-Franche-Comté** | 0.5 | 0.2 | 11 |
| **Bretagne** | 1 | 1 | 2.5 |
| **Centre-Val de Loire** | 1 | 0.5 | 6.5 |
| **Grand Est** | 1 | 0.5 | 5 |
| **Hauts-de-France** | 1 | 0.5 | 7 |
| **Ile-de-France** | 1 | 1 | 3 |
| **Normandie** | 1 | 0.33 | 6 |
| **Nouvelle-Aquitaine** | 1 | 0.5 | 6.5 |
| **Occitanie** | 1 | 1 | 3 |
| **Pays de la Loire** | 1 | 0.5 | 6.5 |
| **Provence-Alpes-Côte d'Azur** | 1 | 0.66 | 6.5 |
| **Total mean** | 0.92 | 0.58 | 5.88 |

Table 2 compares the performance of our sick leave model with the Sentinelles surveillance system. We observe a mean sensitivity of 0.89 (range 0.64–1) and a mean specificity of 0.86 (range 0.80–0.95). When both alerts match, the sick leave model alert was always triggered earlier than the Sentinelles model, with an average lead time of 2.5 weeks (range 0.5–4).

**Table 2:** Performance of the sick-leave model compared to the Sentinelles model: sensitivity, specificity and detection time before ILI peak. For each region, the value of these indicators are averaged over the two years evaluated.

| **Region** | **Sensitivity** | **Specificity** | **Detection time (weeks) before ILI peak** |
| --- | --- | --- | --- |
| **Auvergne-Rhône-Alpes** | 0.87 | 0.81 | 1.5 |
| **Bourgogne-Franche-Comté** | 0.64 | 0.80 | 3 |
| **Bretagne** | 1 | 0.89 | 3 |
| **Centre-Val de Loire** | 1 | 0.83 | 4 |
| **Grand Est** | 0.91 | 0.84 | 4 |
| **Hauts-de-France** | 0.83 | 0.84 | 3 |
| **Ile-de-France** | 0.82 | 0.95 | 1 |
| **Normandie** | 0.93 | 0.80 | 1.5 |
| **Nouvelle-Aquitaine** | 1 | 0.85 | 4 |
| **Occitanie** | 0.79 | 0.95 | 1.5 |
| **Pays de la Loire** | 0.89 | 0.87 | 0.5 |
| **Provence-Alpes-Côte d'Azur** | 1 | 0.87 | 3 |
| **Total mean** | 0.89 | 0.86 | 2.5 |

### 4. Discussion

Workplace absenteeism data can be used by public health surveillance systems to detect emerging infectious disease epidemics (15). Despite this, to this date, few health authorities worldwide use absenteeism data to inform outbreak surveillance. Here, we assessed the potential of workplace absenteeism data to monitor influenza and detect epidemics in France, using an adapted statistical method to analyze this data. We applied this method to a comprehensive national database of workplace absenteeism and validated it against the French national surveillance system based on sentinel GPs. Our results suggest that a system based on workplace absenteeism could be highly sensitive and detect influenza epidemics earlier than the current French surveillance system.

We found that the surveillance system we propose would be able to detect outbreaks 5.9 weeks before the peak and about 2.5 weeks before the Sentinelles system. This suggests that sick-leave data could be almost as timely as emergency visits data that has a timeliness of 3 weeks compared to ILI data systems (16). Our findings in this regard are in line with previously published studies in several contexts worldwide. In a French study from 1994, sick-leave data from a large company allowed to detect flu epidemics with up to 2 weeks of advance (9). In a more recent Belgian study, worker absenteeism data from the Belgian Medical Expertise and from the Belgian railway system was shown to start rising 2–3 weeks in advance and to peak 2 weeks in advance, as compared with ILI data from the Belgian sentinel GP surveillance system (6). In the UK, data on workplace absenteeism among employees of Transport for London peaked up to 2 weeks before the NHS ILI surveillance data(7); and monitoring workplace absence due to “cold”, “cough” or “influenza” among the staff of a large hospital organization was shown to allow the detection of flu epidemics with a significant advance of up to 9 weeks (17).

Very few of these previously published studies included an assessment of the sensitivity and specificity of an absenteeism-based surveillance system. However, in the French study from 1994, the sensitivity and specificity of surveillance based on sick-leave data from a large company were estimated at 74% and 67% for the identification of epidemic weeks and 67% and 94% for the detection of epidemics, with an 80% positive predictive value (9). The UK study based on hospital staff absenteeism also noted that the resulting system did not lead to more false positives than the NHS surveillance data in London.

The quality of the developed model depends strongly on the quality of the data collected within companies. Sick-leave data have the advantage of describing quasi-real individual behavior regarding sick-leave and presence at the workplace. These data are in fact used to enter employees’ pay and are subsequently fulfilled by obligation in the computer system. Our data may not be representative of French population because it describes data from a health-insurer. For instance, it insures few construction companies because they have their own specific insurer. The data also do not include unemployed people. However, representativeness is not necessarily required to build up an outbreak detection system. In fact, outbreak detection aims to detect any unusual expected number of cases to generate signal alarms. Similarly, the GP Sentinelles network does not include all GPs but a small subset of the same practitioners over time.

Another downside of our data is related to the definition of the sick leave rate. The denominator of this rate is the number of workers and it includes workers that are on holiday. The sick leave rate therefore drops during school holidays and the systems do not detect any alerts during those holidays even if they are included in the statistical model as covariates. This is an issue if the epidemic occurs during the holidays and this is the case during the year 2016–2018 where the peak occurs during the Christmas break for two regions. The model could then be more sensitive if the denominator was the number of employees actually at work.

Furthermore, the model accuracy estimated by the false positive rate is strongly related to the definition of cases to detect. The time series of cases is based only on flu cases. However, other epidemics such as gastroenteritis can influence sick leaves data and could be considered for future works: the sick leaves outbreaks may correspond to other disease and may explain the poor specificity in some cases. The detection algorithm could also be improved if the estimation of expected cases could adjust for any potential past exogenous environmental factors such as a terrorist attack, strikes or unexpected bad weather episodes. The model accuracy is moreover strongly related to the method chosen for the algorithm. The Serfling method may actually not be the more accurate model and was chosen to be consistent with the Sentinelles method. Some other regression-based models are known to be more specific, like the Farrington algorithm (18,19).

Another limitation of our study is linked to the fact that we relied on data that were consolidated on an annual basis, and not in real-time. As such, the system is able to detect outbreak based on the dates of sick-leaves, but only retrospectively. A proper integration of sick-leave data into a health surveillance system would thus require an effort to ensure sick-leave data are consolidated and made available in real time. Our results suggest that the resulting increased timeliness of a surveillance system including this data stream may justify this effort. Moreover, these data sometimes already exist in near-real time because of local legislation. For instance, French companies must declare sick leave within 5 days.

Many of the previously published studies assessing the potential of workplace absenteeism data for flu surveillance simply provided visual analyses comparing and correlating absenteeism data with ILI surveillance data (6,7). By contrast, in this work, we propose a statistical approach and algorithm to analyze the French workplace absenteeism data, and to raise alarms when outbreaks are detected. This allows us to both propose a complete surveillance system that could be used in practice provided the data is available, and to fully assess the performance of this surveillance system.

## Data Availability

Influenza-like ilness data from Sentinelles are available online: https://websenti.u707.jussieu.fr/sentiweb/

## Acknowledgement

TD is supported by Association Nationale de la Recherche et de la Technologie and Malakoff Humanis.

JB is supported by the INCEPTION project (PIA/ANR-16-CONV-0005)

PAA is supported by INSERM-ANRS (France Recherche Nord & Sud Sida-HIV Hépatites), grant number ANRS-12377 B104

DS is supported by a Canadian Institutes of Health Research Doctoral Foreign Study Award (Funding Reference Number 164263) as well as the French government through its National Research Agency project SPHINX-17-CE36-0008-01.

## Authors’ contribution

TD, JB, RL: analyzed and extracted the data. TD, JB, PAAT, WD, KJ, LT, MH: Wrote the first draft of the paper. TD, JB, PAAT, RA, OSD, AD, JPG, NN, HN, DRMS, WD, KJ, LT, MNH: designed and discussed statistical analysis and interpreted data. All authors: Read, reviewed and approved the final manuscript.

## Supplementary material

**Figure S1:**
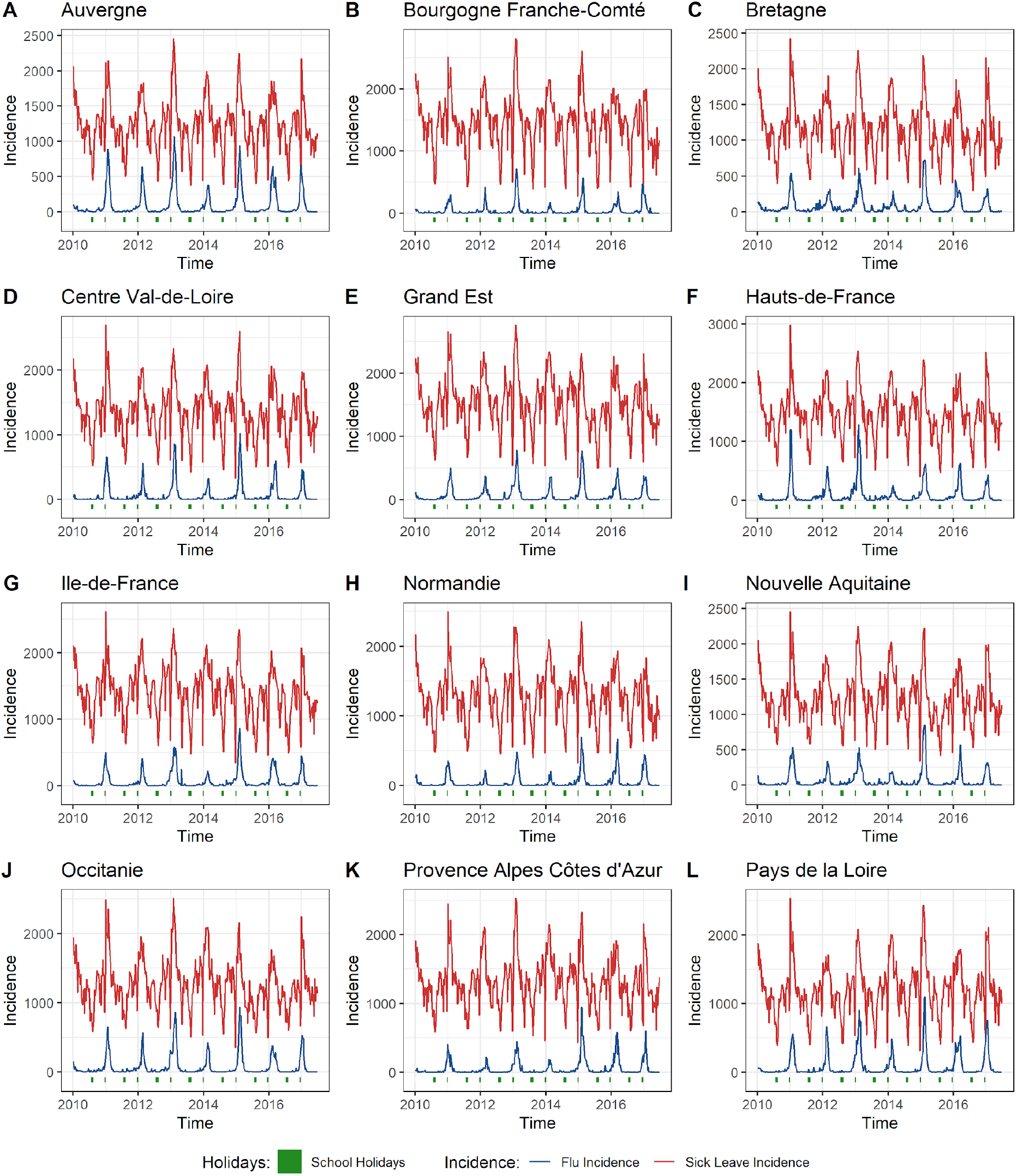
Incidence per 100,000 per week of influenza-like illness and sick leave in twelve French regions, 2010–2017. The Christmas and summer school holidays (increased worker leave periods) are shown at the bottom.

**Figure S2:**
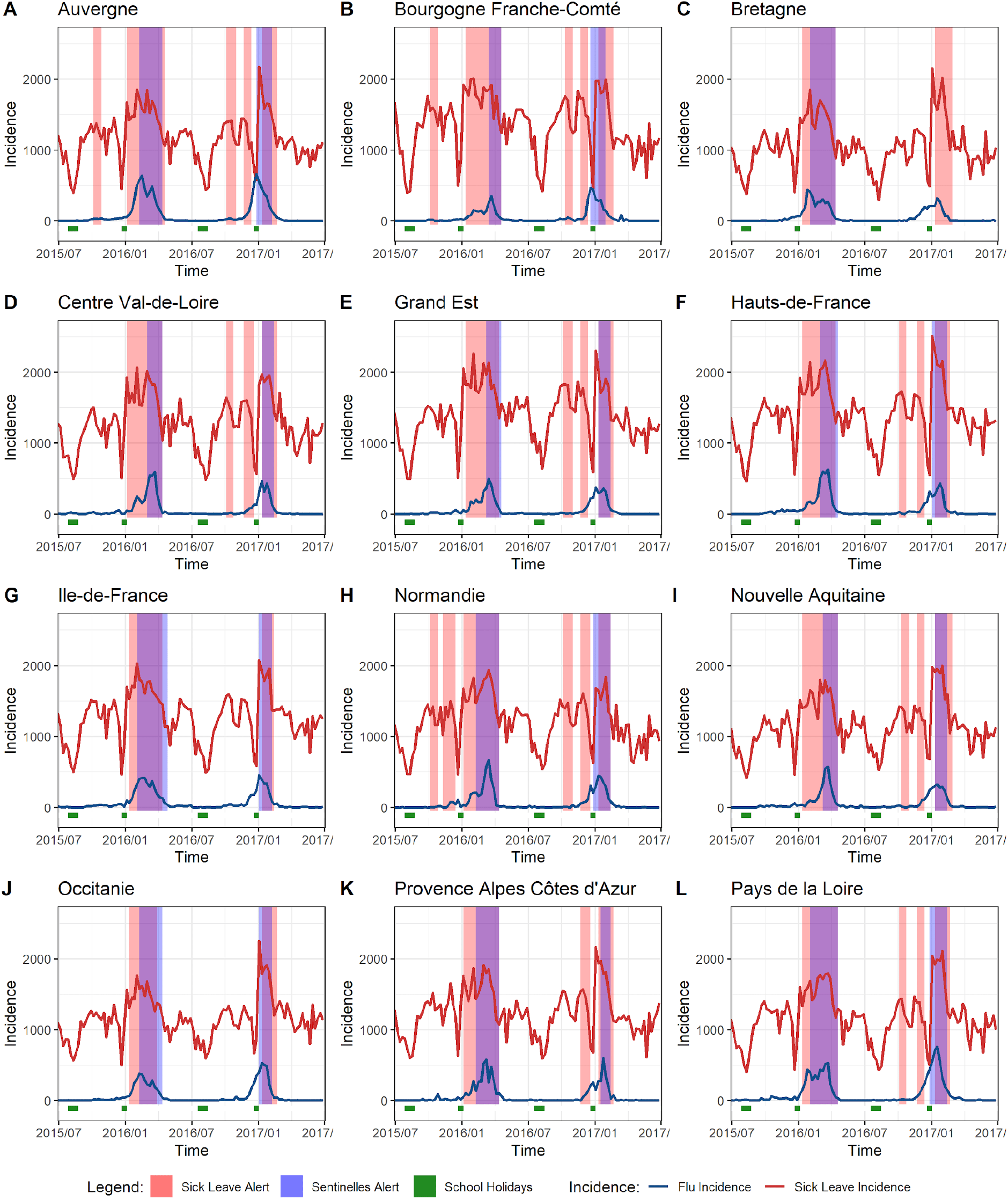
Incidence of influenza-like illness and sick leave, 2015–2017, and alerts from the Sentinelles and the sick-leave models, in twelve French regions. The Christmas and summer school holidays (increased worker leave periods) are shown at the bottom.

